# Complement activity in myasthenia gravis is independent of autoantibody titer and disease severity

**DOI:** 10.1101/2021.01.15.21249875

**Authors:** Miriam L. Fichtner, Michelle D. Hoarty, Douangsone D. Vadysirisack, Richard J. Nowak, Kevin C. O’Connor

## Abstract

Acetylcholine receptor (AChR) autoantibodies, found in patients with autoimmune myasthenia gravis (MG), can directly contribute to disease pathology through activation of the classical complement pathway. Accordingly, complement inhibitors are used as a therapeutic strategy, but the response can be heterogeneous even though AChR autoantibodies are present. The mechanisms underlying the variable response are not defined. Yet there is a need for further understanding so that responses can be better predicted. There is a broad spectrum of circulating complement activity levels activity among MG patients. It is not clear whether this activity associates with disease burden or the circulating levels of autoantibodies. We measured complement activity and investigated these associations in MG patients as a means to explore candidate biomarkers. Most study subjects had complement activity within the range defined by healthy controls and no association between this activity and disease burden or AChR autoantibody titer was observed. Assays measuring the complement activating properties of AChR autoantibodies are needed to identifying patients expected to respond to complement inhibitor-based treatments.

## Introduction

The most common subtype of autoimmune myasthenia gravis (MG) is characterized by pathogenic autoantibodies targeting the nicotinic acetylcholine receptor (AChR). These autoantibodies directly contribute to disease pathology primarily, though not exclusively, through activation of complement. Eculizumab, a therapeutic complement inhibitor, was demonstrated to be effective in a phase 3 trial with subsequent approval for its use in treating acetylcholine receptor antibody-positive MG [1-3].Clinical response to this treatment can be heterogeneous; many patients respond well or show a delayed response while, others respond poorly. The mechanisms underlying the variable response are not known. However, there is a need for further understanding so that responses can be better predicted. While the majority of MG patients have circulating complement activity levels that are within the normal range, there is a broad spectrum of such activity among MG patients. It is not clear whether this activity associates with MG disease burden or the circulating levels of autoantibodies. We measured complement activity and investigated associations with disease burden and autoantibody titers in MG patients as a means to explore candidate biomarkers.

## Materials and methods

This study was approved by the Human Investigation Committee at the Yale School of Medicine (clinicaltrials.gov || NCT03792659). Informed consent was obtained from all participants. Peripheral blood was collected from AChR MG and healthy control (HC) participants. All MG participants met definitive diagnostic criteria for MG, including positive serology. Treatment status of the MG patient cohort (N=51) was heterogeneous: no current therapy (N=4), immunotherapy naïve (N=22), cholinesterase inhibitor (N=10), corticosteroids (N=11), both corticosteroids and azathioprine (N=1), both corticosteroids and cholinesterase inhibitor (N=1), IVIG (N=1), or plasmapheresis (N=1). Antibody-sensitized sheep erythrocytes (Complement Tech) were used to determine the hemolytic activity of the classical complement pathway (CH50 units) [4]. Statistical calculations were performed with Prism Software (GraphPad; version 8.0).

## Results

We tested 51 AChR MG participants (mean age: 59.3 +/− 18.8 yrs) and 20 healthy controls (HC) (mean age: 34.4 +/− 13.3 yrs) for complement activity by hemolytic CH50 assay. The majority of AChR MG samples had complement activity (CH50 values) within the range defined by the HDs **(Figure 1A)**, while several patients had reduced complement activity and several others had elevated activity **(Figure 1A)**. No association (ns; p=0.8) between disease burden, as measured by the MG Composite score, and complement activity was observed **(Figure 1B)**. Similarly, no significant (ns; p=0.5) correlation between complement activity and AChR autoantibody titer was observed **(Figure 1C)**.

**Figure 1.**
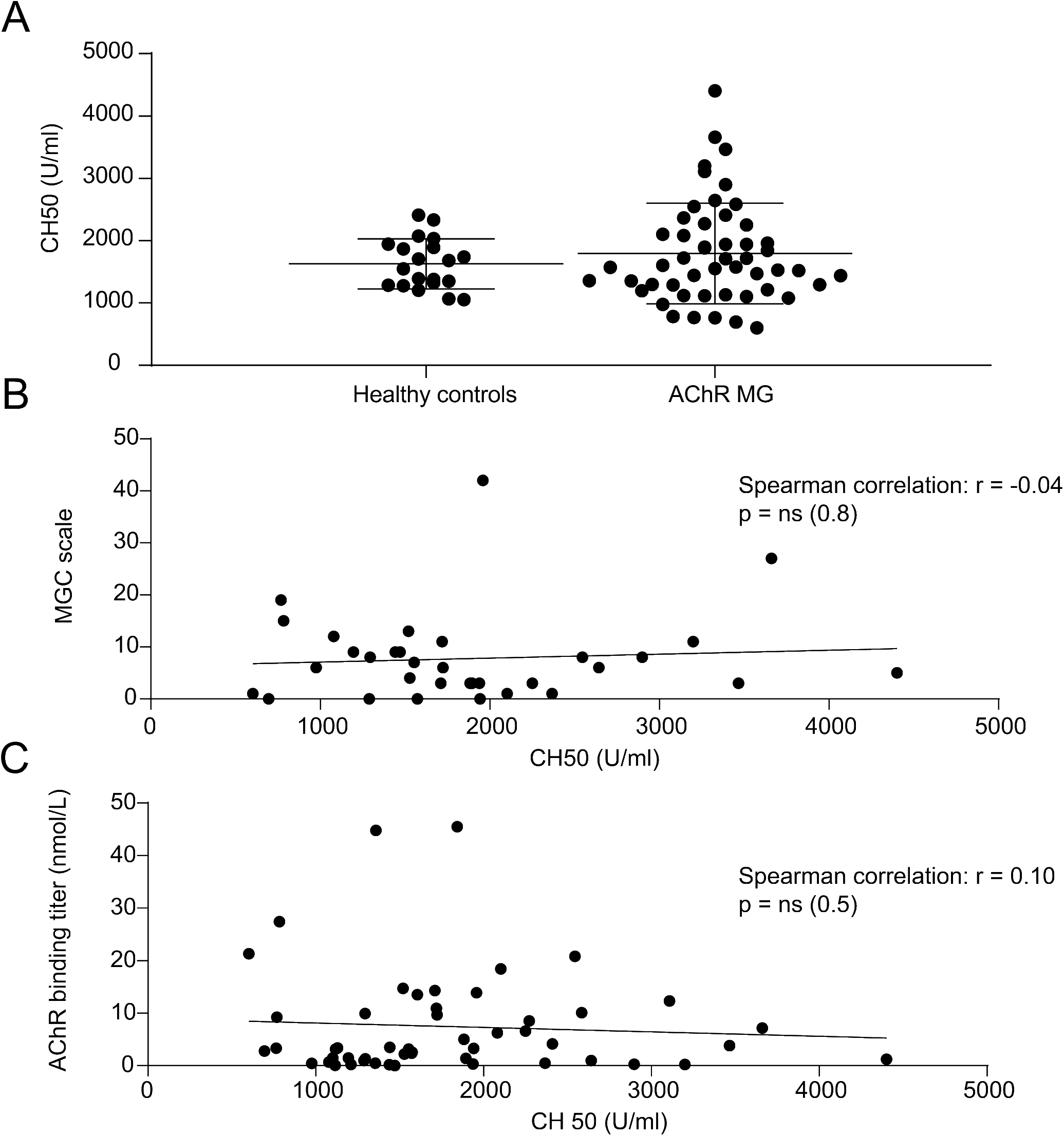
Complement activity in AChR MG does not associate with clinical status or autoantibody titer. Complement activity in the serum of AChR MG participants was measured by CH50 hemolysis assay. The assay measures the activation of the classical complement pathway by testing the ability of the complement components of sera samples to lyse antibody-sensitized sheep erythrocytes. (A) AChR MG participants patients (N=51) and HC (N=30) were screened by CH50 hemolysis assay. (B) Correlation of complement activity to the Myasthenia Gravis Composite (MGC) score. The MGC score values were available for 33 out of the 51 participants. (C) Correlation of complement activity to AChR titer (N=51). The linear regression is shown with Spearman correlation values.

## Discussion

This study suggests that complement activity in most AChR MG patients is likely normal and that there is no correlation of complement activity to AChR autoantibody titer or disease severity/burden. The assays that are routinely used to diagnose MG, measure the capability of the polyclonal serum derived autoantibodies to bind to the AChR, but not their pathogenic properties. Three major pathogenic mechanisms have been identified for AChR autoantibodies. The first describes AChR autoantibodies blocking the access of acetylcholine to the AChR and thus hindering the neuromuscular signal transduction [6]. The second, termed antigen modulation, describes AChR receptor crosslinking followed by receptor internalization resulting in a reduced number of available receptors [7]. The third involves activation of the classical complement pathway by AChR autoantibodies [8]. There may also be AChR autoantibodies that bind the receptor but have no pathogenic capacity. This complex heterogeneity of circulating AChR autoantibody properties may contribute to the poor correlation between AChR autoantibody titer and disease severity [9]. This study indicates that there is additionally a poor correlation between complement activity and both AChR autoantibody titer and disease burden in AChR MG patients. Currently, there is no established approach for measuring the complement activating properties of AChR autoantibodies. Such a tool would be valuable for identifying patients expected to respond to complement inhibitor-based treatments in an effort toward applying precision medicine.

## Data Availability

The data reported in this paper is available upon request.

## Acknowledgements

The authors thank Bailey Munroe Sheldon who helped with the collection of the clinical data. The authors thank Karen Boss for providing editorial assistance.

